# Accounting for age-related increases in HbA1c more accurately quantifies risk of Type 1 Diabetes progression in islet autoantibody-positive adults

**DOI:** 10.64898/2026.02.19.26346463

**Authors:** Erin L. Templeman, Nick Thomas, Susan Martin, Diane K. Wherett, Maria J. Redondo, Jennifer Sherr, Alessandra Petrelli, Laura M. Jacobsen, Falastin Salami, Jacqueline Lonier, Carmella Evans Molina, Jay Sosenko, Inês Barroso, Richard A. Oram, Emily K. Sims, Lauric A. Ferrat

## Abstract

**Objective:** HbA1c thresholds used to define dysglycemia in autoantibody-positive individuals at risk for type 1 diabetes do not account for age-related increases in HbA1c and may overestimate progression risk in adults. We evaluated whether age-adjusted HbA1c or a higher HbA1c threshold improves risk stratification across age groups.

**Research Design and Methods:** We analyzed 5,024 autoantibody-positive relatives (3,720 children and 1,304 adults) participating in the TrialNet Pathway to Prevention study. Age-related HbA1c effects were modelled using 6,273 adults from the population-based Exeter 10,000 cohort. Progression risk was compared using the standard dysglycemia threshold (HbA1c ≥ 5.7% [39 mmol/mol]), age-adjusted HbA1c, and an alternative threshold of HbA1c ≥6.0% (42 mmol/mol).

**Results:** Using HbA1c ≥ 5.7%, children had higher 1-year progression risk than adults among single autoantibody–positive participants (38% [95% CI 28, 47] vs. 13% [7.2, 19]) and multiple autoantibody–positive participants (55% [49, 60] vs. 38% [27, 47]; both p<0.001). Age adjustment reduced these differences; progression risk was similar among single autoantibody–positive participants (38% [28, 47] vs. 27% [13, 39]; p=0.32), with attenuated differences among multiple autoantibody–positive participants. An HbA1c threshold ≥6.0% yielded comparable progression risk between adults and children across autoantibody subgroups. In post hoc analyses, adults aged <30 years had progression risk similar to children (p=0.1).

**Conclusions:** Age-related variation in HbA1c influences dysglycemia classification in adults at risk for type 1 diabetes. Age-adjusted HbA1c or a higher HbA1c threshold (≥6.0% [42 mmol/mol]) in adults ≥30 years identifies individuals with progression risk comparable to children and may improve age-specific risk stratification in prevention seungs.

## Introduction

Approval of teplizumab to delay clinical type 1 diabetes onset has yielded a worldwide expansion in type 1 diabetes screening (1,2). This expansion creates a growing need for simple, reproducible markers to identify islet autoantibody (AB) positive individuals at highest risk of progression to guide timely follow-up and therapy initiation. In islet AB positive individuals, elevated glucose values within the non-diabetes range are a well-established predictor of progression to clinical or stage 3 type 1 diabetes (3). In fact, progression to stage 2 disease is defined by the presence of multiple ABs plus development of dysglycaemia, which can be defined using oral glucose tolerance testing (OGTT), fasting glucose, or HbA1c (≥ 5.7% or 39 mmol/mol) (4). Because stage 2 disease is required for eligibility for clinically available disease-modifying therapy, accurate identification carries substantial clinical implications. Among measures used to define stage 2 disease, HbA1c is widely used in clinical care given that it is convenient, can be obtained without fasting or glucose stimulation, imposes minimal burden on patients, and is familiar to clinicians.

A key limitation of HbA1c is that it reflects both glycaemic exposure and a range of non-glycaemic influences, such as haemoglobin variants, variation in erythrocyte lifespan, ancestry-associated differences, as well as age-related non-glycaemic changes (modest slowing of red blood cell turnover, shivs in erythropoiesis, and increased intrinsic glycation rates) that occur independent of glucose (5– 7). These factors contribute to higher HbA1c values with increasing age in the general population, a pattern that may differ from trajectories in a selected cohort with a well-defined risk of type 1 diabetes (5–7). Specifically, in the general population without diabetes, mean HbA1c is lower in children than in adults. Within children in the general population mean HbA1c is approximately 32 mmol/mol (5.1%), with only 0.15% of children in the general population having an HbA1c above the threshold used to define dysglycaemia in stage 2 type 1 diabetes (8–11). A much higher proportion of healthy adults (approximately 30%) in the general population have an HbA1c over the dysglycaemic threshold, a prevalence about 200-fold higher compared to children (12). These striking age-related differences suggest that an HbA1c ≥ 5.7% (39 mmol/mol) is far more likely to reflect type 1 diabetes associated disease progression in AB positive children compared to adults. Therefore, higher thresholds may be required to identify true progression of type 1 diabetes in older age groups. Notably, glucose thresholds for defining elevated risk of type 2 diabetes (“pre-diabetes”) vary globally, with the American Diabetes Association recommending HbA1c ≥ 5.7% (39 mmol/mol), while both the International Expert Committee (IEC) and World Health Organization (WHO) suggest HbA1c ≥ 6.0% (42 mmol/mol)(13,14).

We aimed to evaluate HbA1c as a predictor of progression to stage 3 type 1 diabetes in a large cohort of AB positive adults, and to assess whether adult-specific approaches to the HbA1c-based definition of dysglycaemia might lessen observed differences in progression risk between adults and children as compared to applying one single HbA1c threshold to both age groups.

## Methods

### Participants

We evaluated progression risk in single and multiple islet AB positive adults and children from the TrialNet Pathway to Prevention cohort (ClinicalTrials.gov ID: NCT00097292) meeting stage 2 type 1 diabetes based on HbA1c cutoffs for stage 2 disease (HbA1c ≥ 5.7% or 39 mmol/mol) (15). Pathway to Prevention participants were followed with serial OGTT and HbA1c monitoring until diagnosis of stage 3 type 1 diabetes or enrolment in a prevention trial, with the last included visit before April 23, 2023. Stage 3 type 1 diabetes was defined by standard glycaemic criteria (fasting plasma glucose ≥ 7.0 mmol/L or ≥ 126 mg/dL, 2-h plasma glucose ≥ 11.1 mmol/L or ≥ 200 mg/dL, or HbA1c ≥ 6.5% or ≥ 48 mmol/mol) (4). Participants with stage 3 type 1 diabetes at screening, no HbA1c data, or no follow-up visits were excluded (Supplementary Figure 1). The detailed TrialNet protocol has been published previously (15).

We also used general population data from Exeter 10,000 (EXTEND) to age-adjust HbA1c in TrialNet to evaluate the impact of age on progression risk. EXTEND is a cross-sectional population-based cohort recruited from the community and primary care clinics in the South West UK (16). For this analysis we excluded participants in EXTEND without baseline HbA1c, with diabetes, or with outlier HbA1c measurements (> 4 standard deviations from the mean). Participants were selected in the same age-range (18 to 58 years) as adults in the TrialNet Pathway to Prevention (Supplementary Figure 2).

### HbA1c measurement

HbA1c was measured in both TrialNet and EXTEND using ion-exchange high-performance liquid chromatography on the TOSOH G8 platform, with TrialNet values standardized to the NGSP/DCCT reference method (reported as %) and EXTEND values calibrated to the IFCC reference preparation (reported as mmol/mol).

### Autoantibody measurement

TrialNet evaluated ABs to glutamic acid decarboxylase (GAD), insulin (micro-insulin antibody [mIAA] assay), insulinoma-associated antigen-2 (IA-2), and zinc transporter-8 (ZnT8), using radiobinding biochemical assays participating in the Islet Autoantibody Standardization Program (17,18). Detailed information on the protocol of AB measurement is available (15).

### Statistical Analysis

Time-to-event analyses were conducted to evaluate progression risk to stage 3 type 1 diabetes, with dysglycaemia (defined by HbA1c) as the starting point. Dysglycaemia was defined both from OGTT data at initial staging and from within follow-up/monitoring visits. Kaplan-Meier survival curves were generated to estimate the risk of stage 3 type 1 diabetes from the point of a dysglycaemic HbA1c. Survival distributions between groups were compared using log-rank tests.

To adjust for the effect of age on HbA1c, a linear regression model was developed in the EXTEND dataset, with age as the predictor, to estimate the expected HbA1c values for ages 18-58 years (the age range in included adult TrialNet participants). Adjusted HbA1c values for TrialNet participants were then derived by removing the estimated age-related effect from the observed HbA1c values. A detailed description of the age adjustment procedure is provided in the Supplementary Methods.

We also evaluated two other approaches by adjusting the HbA1c threshold for adults to match the risk of disease progression observed in children using HbA1c ≥ 5.7% (3ti mmol/mol) cutoff for dysglycaemia. A threshold of HbA1c ≥ 6.0% (42 mmol/mol) was selected based on guidelines for type 2 diabetes in some countries (e.g., UK NICE, Australian and WHO-aligned recommendations)(19),

1. **First pragmatic approach:** Applied a uniform dysglycaemic threshold of 42 mmol/mol for all adults.
2. **Second pragmatic approach:** Set the threshold at 42 mmol/mol for adults aged 30 years or older, derived from previous literature (20,21), accounting for known differences in disease progression in younger adults.

All statistical analyses were computed with R version 4.4.0 (2024-04-24) with the package *survival*. Statistical significance was defined as a two-sided p-value < 0.05.

## Results

Of 5,024 relatives followed in the TrialNet Pathway to Prevention, 3,720 (74%) were children (<18 years at screening; followed for median [IQR] of 3.74 years [1.65, 6.94]), and 1,304 (26%) were adults (≥18 years old at screening; followed for 3.46 years [1.55, 6.07]), characteristics shown in Supplementary Table 1. Prevalence of a dysglycaemic HbA1c ≥ 5.7% (39 mmol/mol) at screening was lower in children compared to adults (5.3% vs 8.4%, respectively, p<0.001). Of the 1,304 adults screening AB positive, 990 (76%, 990/1304) were single AB positive and 314 (24%, 314 /1304) were multiple AB positive. As expected, prevalence of dysglycaemic HbA1c at screening was higher in adults with multiple ABs compared to single AB positive adults (12.7% vs 7.1%, respectively, p<0.001). Among single AB positive individuals, dysglycaemic HbA1c was more common in adults than in children (7.1% vs 3.6%, p=0.03). The characteristics of the adult population stratified by AB number are displayed in Supplementary Table 2. Within adult non-progressors, as predicted from literature, HbA1c increased with age (coef: 0.0134 mmol/mol per year [95%CI 0.063, 0.116]; p<0.001; Supplementary Figure 3).

### Children with dysglycaemic HbA1c were more likely to progress to stage 3 type 1 diabetes compared to adults

We next compared absolute progression risk amongst children and adults with dysglycaemic HbA1c values (HbA1c ≥ 5.7% or 39 mmol/mol). Here, the 1-year risk of progression to type 1 diabetes was dramatically higher in children than in adults. The risks for participants positive for a single AB were nearly three-fold higher for children compared to adults (children: 38% [95%CI 28, 47]; adults: 13% [95%CI 7.2, 19]; p<0.001; Figure 1A). For those positive for multiple ABs, the estimated risk was 55% [95%CI 49, 60] in children and 38% [95%CI 27, 47] in adults (p<0.001; Figure 1B).

**Figure 1:**
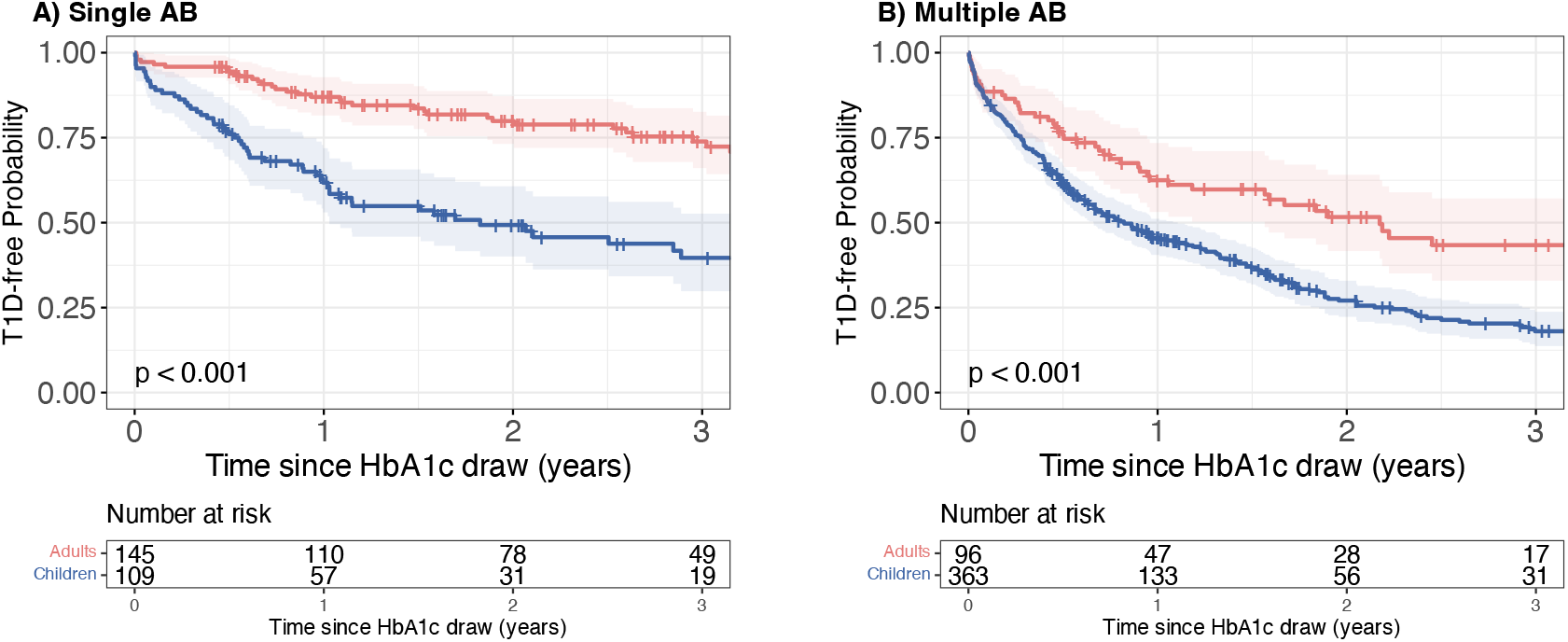
Kaplan-Meier survival curves of TrialNet participants stratified by age (Adults [red] vs. children [blue]) from the time of dysglycaemia. Dysglycaemia was defined as HbA1c ≥ 5.7% (39 mmol/mol). Survival represents time from dysglycaemia to clinical type 1 diabetes. Panel A: Participants who were positive for a single autoantibody at the time dysglycaemia was identified using HbA1c ≥ 5.7% (39 mmol/mol). Panel B: Participants who were positive for multiple autoantibodies at the time dysglycaemia was identified using HbA1c ≥ 5.7% (39 mmol/mol). All survival curves are truncated when fewer than ten participants remain under follow-up.

### Accounting for rising HbA1c with age gives closer risk estimates for adults compared to children

Given discrepancies in absolute risk of diabetes progression between adults and children who meet dysglycaemic HbA1c cutoffs, we next examined whether age-adjusted HbA1c values would yield similar progression risks for both groups meeting the 39 mmol/mol (5.7%) threshold. First, to generate age-adjusted values we considered an external dataset from participants without diabetes in the EXTEND study (see Supplementary Table 3 for study characteristics). The age range is similar between the two cohorts, however the lower number of TrialNet adults (n = 336 screened over 30 years old) results in a reduced mean age in the TrialNet cohort compared to the EXTEND cohort (p<0.001). Analysis of the EXTEND study confirmed a significant effect of age on HbA1c levels, with a coefficient of 0.135 [95%CI 0.125, 0.145] (p-value < 0.001; Figure 2), i.e. for each decade of life, HbA1c increases by ∼1 mmol/mol or 0.1%. To account for this age-related variation, we utilized the age effect coefficient (0.135) to age-adjust HbA1c levels to an equivalent value in an 18-year-old. For instance, for a 40-year-old adult with a measured HbA1c of 6.0% (42 mmol/mol), the age-adjusted HbA1c dysglycaemia threshold would be HbA1c ≥ 5.7% (39 mmol/mol) (Supplementary Table 4 contains further examples; values are rounded to aid clinical interpretation).

**Figure 2:**
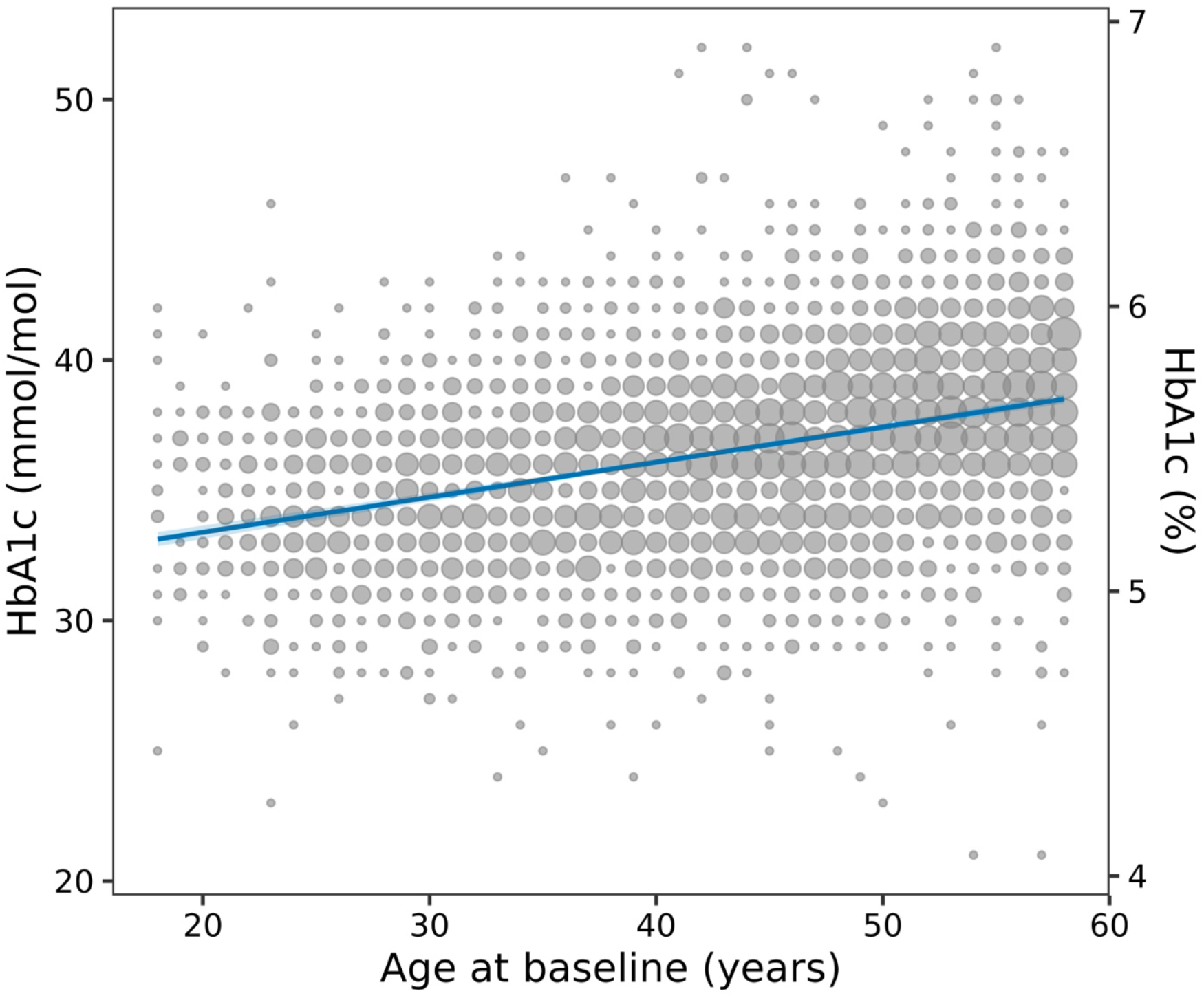
Linear Regression model and scatter plot of HbA1c values (mmol/mol on left, % on right) by age (years) in EXTEND. Each point represents one or more participants with the same age and HbA1c value; the size of the point is proportional to the number of individuals at that exact combination. The solid blue line indicates the fitted linear regression model and the shaded band around the regression line represents the 95% confidence interval. The coefficient or effect of age when predicting HbA1c (mmol/mol) is 0.135 (95% CI [0.125, 0.145]. For each decade of life, HbA1c (mmol/mol) increases by around 1 mmol/mol or 0.1%.

Using the age-adjusted HbA1c with a threshold of HbA1c ≥ 5.7% (39 mmol/mol) for defining dysglycaemia reduced the number of adults meeting the measured HbA1c cutoff for dysglycaemia (n=105 meeting age-adjusted HbA1c ≥ 39 mmol/mol vs n=241 with measured HbA1c ≥ 39 mmol/mol). Using the age-adjusted HbA1c threshold produced similar 1-year risk between single AB positive children and adults (children: 38% [28, 47]; adults: 27% [13, 39]; p=0.32; Figure 3A). In multiple AB positive participants, using the age-adjusted HbA1c reduced the difference in 1-year risk between multiple AB positive children and adults (children: 55% [49, 60]; adults: 45% [29, 58]; p=0.03; Figure 3B).

**Figure 3:**
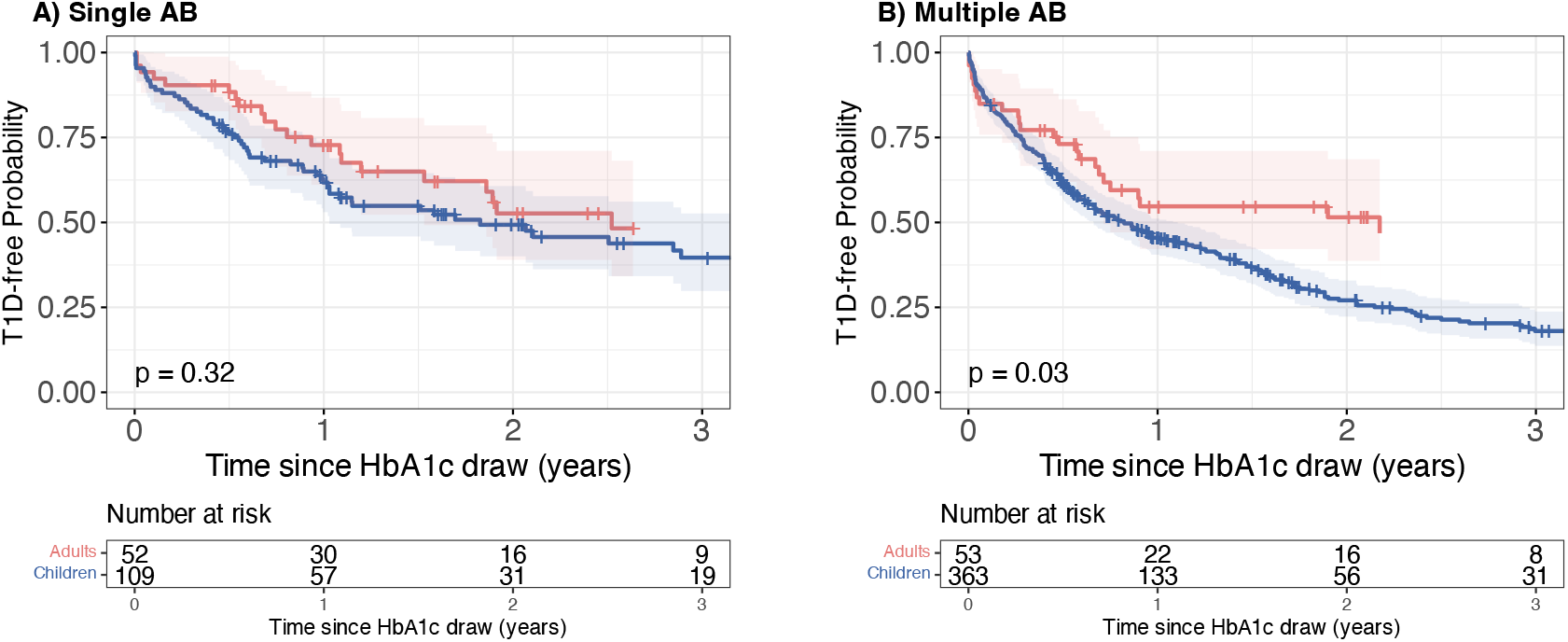
Kaplan-Meier survival curves of TrialNet participants stratified by age (Adults [red] vs. children [blue]) from the time of dysglycaemia. Dysglycaemia was defined as age-adjusted HbA1c ≥ 5.7% (39 mmol/mol). Survival represents time from dysglycaemia to clinical type 1 diabetes. Panel A: Participants who were positive for a single autoantibody at the time dysglycaemia was identified using age-adjusted HbA1c ≥ 5.7% (39 mmol/mol). Panel B: Participants who were positive for multiple autoantibodies at the time dysglycaemia was identified using age-adjusted HbA1c ≥ 5.7% (39 mmol/mol). All survival curves are truncated when fewer than ten participants remain under follow-up.

### Higher HbA1c threshold in adults

We also asked if a pragmatic approach using an increased measured HbA1c threshold cutoff to identify adults with dysglycaemic HbA1c would yield a similar risk of disease progression to children with the HbA1c ≥ 5.7% (39 mmol/mol) threshold for defining dysglycaemic HbA1c. Here we applied the WHO pre-diabetes threshold of HbA1c ≥ 6.0% (42 mmol/mol) to define dysglycaemia in adults, and HbA1c ≥ 5.7% (39 mmol/mol) in children. Using the higher HbA1c threshold of HbA1c ≥ 6.0% (42 mmol/mol) to define dysglycaemia reduced the number of adults meeting dysglycaemia criteria (n=86 meeting higher threshold HbA1c ≥ 6.0% or 42 mmol/mol vs n=241 with measured HbA1c ≥ 5.7% or 39 mmol/mol). Using this higher threshold for adults, there was no significant difference in progression risk in single AB positive children compared to adults (children: 38% [28, 47]; adults: 22% [9,33]; p=0.07; Figure 4A). Similarly, there was no difference between multiple AB positive children and adults (children: 55% [49,60]; adults: 44% [23,59]; p=0.3; Figure 4B).

**Figure 4:**
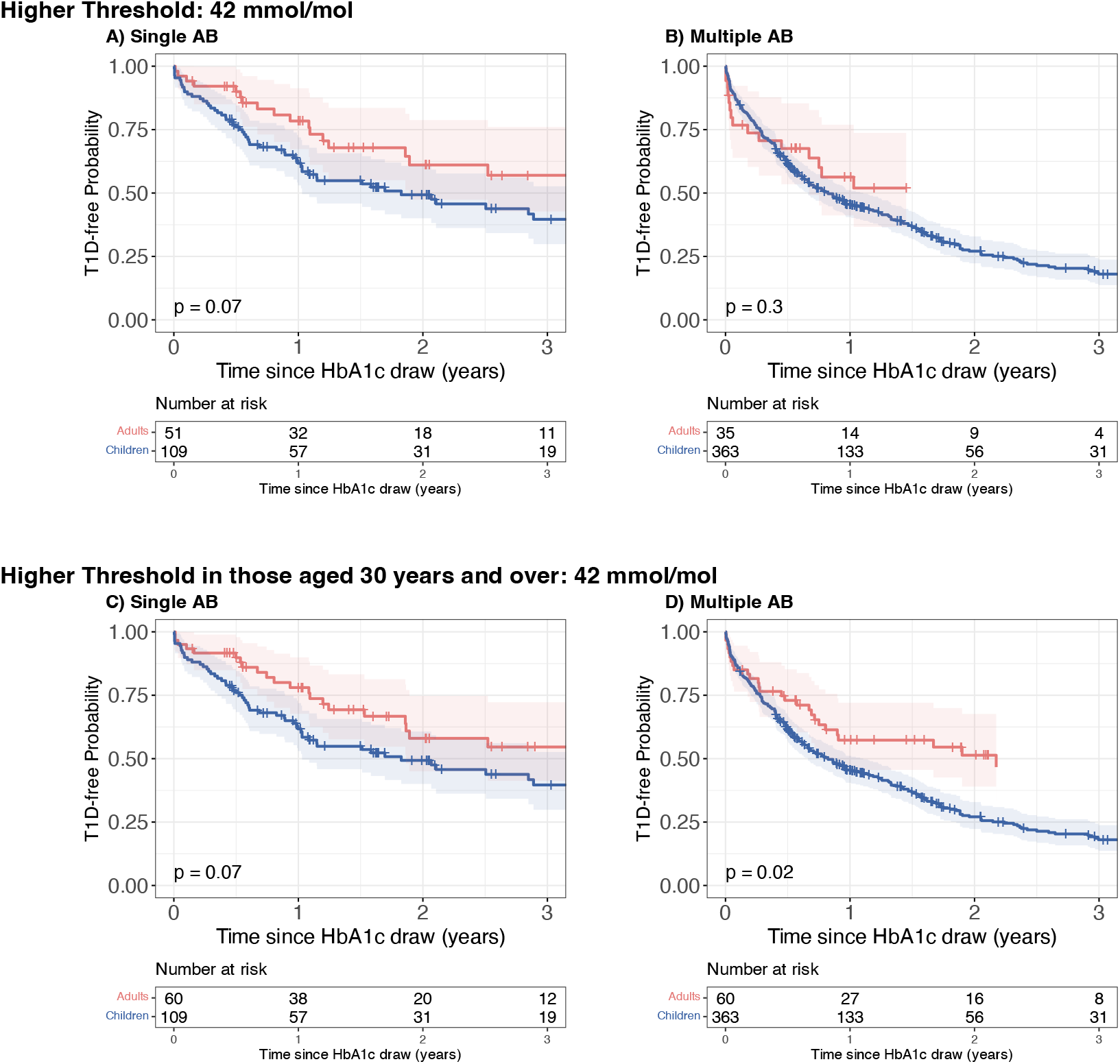
Kaplan–Meier survival curves of TrialNet participants stratified by age (Adults [red] vs. Children [blue]) at the time of dysglycaemia. Survival represents time from dysglycaemia to clinical type 1 diabetes. Panels A and B (top row): Dysglycaemia defined using an HbA1c threshold of ≥ 6.0% (42 mmol/mol) applied to all adults, with children classified using the original threshold (HbA1c ≥ 5.7% or 39 mmol/mol). Panels C and D (bottom row): Dysglycaemia defined using an HbA1c threshold of ≥ 6.0% (42 mmol/mol) applied to adults aged ≥ 30 years, while adults aged < 30 years and children were classified using the original HbA1c threshold (HbA1c ≥ 5.7% or 39 mmol/mol). Panel A and C: Participants positive for a single autoantibody at the time dysglycaemia was identified under the specified HbA1c criteria. Panel B and D: Participants positive for multiple autoantibodies at the time dysglycaemia was identified under the specified HbA1c criteria. All survival curves are truncated when fewer than ten participants remain under follow-up.

### Higher HbA1c threshold only in those aged over 30 years

As noted above, HbA1c levels rise progressively with age in adults, amplifying the need for age-specific risk assessment. Using the original threshold, HbA1c ≥ 5.7% (39 mmol/mol), we examined if this impacted differently in those aged under 30 years compared to those aged ≥ 30 years (defined using previous literature) (20,21). Adults aged under 30 years had higher type 1 diabetes risk compared to adults aged ≥ 30 (p<0.001); the adults aged under 30 years had comparable risk to children (p=0.12; Supplementary Figure 4).

Based on this, we next tested a pragmatic approach applying the higher HbA1c threshold (HbA1c ≥ 6.0% or 42 mmol/mol) only to adults aged ≥ 30 years, while retaining the HbA1c ≥ 5.7% (39 mmol/mol) threshold for those <30 years. Using this approach yielded comparable results to an age-adjusted HbA1c or a uniform adult threshold, with 1-year risk of progression in dysglycaemic single AB positive (22% [10,32]) compared to children (38% [28,47]; p=0.07; Figure 4C), and in dysglycaemic multiple AB positive adults (43% [28,54]) compared to children (55% [49,60]; p=0.02; Figure 4D).

Direct comparison of adults identified as dysglycaemic showed that risks were comparable when applying the higher threshold for dysglycaemia in all adults or only those ≥30 years (p=0.95; Supplementary Figure 5). Children consistently showed higher risk than adults, irrespective of threshold approach (p<0.001; Supplementary figure 6).

### Comparison of approaches for participants above threshold

We compared whether the age-adjusted HbA1c threshold (HbA1c ≥ 5.7% or 39 mmol/mol) or a higher measured HbA1c threshold (HbA1c ≥ 6.0% or 42 mmol/mol) performed similarly in identifying adults at increased risk of progression. Adults identified using the age-adjusted HbA1c at screening (n=105) had a 1-year type 1 diabetes risk of 36% [26, 45], which was comparable to those identified using the higher measured cutoff (n=86, 30% [19, 40]; p=0.6). Seventy-four adults were identified as high risk by both approaches (86% of those meeting the age-adjusted cutoff and 70% of those meeting the higher measured cutoff). When stratified by AB number, progression risk remained similar between those identified by the age-adjusted and higher measured HbA1c thresholds for both single AB positive adults (p=0.39) and multiple AB positive (p=0.54).

### Comparison between adults and children below dysglycaemia threshold

We also asked if changes in threshold approaches for adults may underestimate progression risk in adults that no longer met more conservative dysglycaemia criteria. Children below the HbA1c threshold of 5.7% (39 mmol/mol) consistently showed higher risks of type 1 diabetes compared with adults below the dysglycaemia threshold across all approaches. Specifically, the 1-year risk among children below this threshold was 6.9% [95% CI 6.1, 7.8], whereas the corresponding risk in adults was substantially lower: 3.4% [95% CI 2.4, 4.4] using the standard threshold. Corresponding risk in adults was 4.0% [95% CI 3.0, 5.1] with the age-adjusted HbA1c approach, 4.4% [95% CI 3.3, 5.5] using the 6.0% (42 mmol/mol) cutoff for all adults, and 3.9% [95% CI 2.9, 4.9] when the 6.0% (42 mmol/mol) threshold was restricted to adults aged 30 years or older.

Within the intermediate HbA1c range of 39–41 mmol/mol (5.7–5.9%; between ADA and WHO thresholds), single AB positive adults had a lower 1-year risk of 6.7% [95% CI 2.1, 11] compared to children with comparable HbA1c values (32% [95%CI 21, 41]; p<0.001; Supplementary Figure 7A). Among multiple AB positive adults within the 5.7-5.9% (39-41 mmol/mol) HbA1c interval, risk was lower than in children with a single AB in the same HbA1c range (adults: 32% [95%CI 20, 42]; children: 45% [95%CI 39, 51]; p=0.001; Supplementary Figure 7B).

When restricting the analysis to young adults (<30 years) with HbA1c between 5.7 and 5.9% (39 - 41 mmol/mol), the 1-year risk of type 1 diabetes the risk profile was similar to that observed in children with HbA1c between 5.7 and 5.9% (adults: 42% [95% CI 36, 47]; children: 36% [95% CI 19, 49]; p=0.1). Overall, these findings indicate that young adults with HbA1c between 5.7 and 5.9% (39 and 41 mmol/mol) have a risk profile similar to that of children in the same HbA1c range, supporting the rationale for maintaining the 5.7% (39 mmol/mol) threshold for adults under 30 years.

## Conclusions

In this cohort of islet AB positive relatives, we observed that, while HbA1c is a clinically valuable predictor of progression to stage 3 type 1 diabetes, its prognostic performance differs substantially between adults and children. At the current proposed ADA dysglycaemia threshold of HbA1c ≥ 5.7% (39 mmol/mol), children progressed substantially more rapidly than adults (p<0.001). This suggests that identification of adults with similar progression risk would require alternative HbA1c criteria defining dysglycaemia in AB positive adults.

Adjusting HbA1c for age, using data from a large non-diabetic population-based adult cohort, reduced differences in progression risks between adults and children, with the greatest alignment observed among single autoantibody positive participants, in which progression risks were no longer statistically different. Among multiple autoantibody positive participants, age adjustment attenuated but, in some cases, did not eliminate the higher progression risk observed in children, consistent with persistent differences in disease tempo beyond age-related effects on HbA1c. These findings suggest that part of the discrepancy at the 5.7% (39 mmol/mol) threshold reflects age-related increases in HbA1c unrelated to type 1 diabetes disease-related activity. In the general population without diabetes, HbA1c ≥ 5.7% (39 mmol/mol) is rare in children but relatively common in adults (12), a pattern driven by both glycaemic and non-glycaemic influences. Consequently, an HbA1c of 5.7% (39 mmol/mol) in an autoantibody-positive child is far more likely to reflect early type 1 diabetes than the same value in an adult.

Several non-glycaemic biological mechanisms contribute to rising HbA1c with age. Glycation rates rise with age at a given glucose concentration, potentially due to changes in protein turnover or intracellular glucose handling, and that modest prolongation of erythrocyte lifespan may also contribute to higher HbA1c independent of mean glycaemia (22,23). These processes help explain why HbA1c values increase across adulthood in the general population and why higher diagnostic thresholds are used internationally for identifying dysglycaemia or “pre-diabetes” in type 2 diabetes (6.0% or ≥ 42 mmol/mol).

Motivated by these age-related differences, we evaluated the WHO threshold, HbA1c ≥ 6.0% (42 mmol/mol), in autoantibody-positive adults. This higher cutoff produced adult progression risks that are more closely aligned with those of children and younger adults at the ADA threshold, HbA1c ≥ 5.7% (39 mmol/mol), offering a simple and clinically practical approach for adult risk stratification. Younger adults (<30 years) demonstrated risks similar to children and did not require a higher threshold, supporting continuity of clinical criteria between pediatric and young adult care.

Among participants positive for multiple ABs, a significant difference in progression risk persisted even aver applying the age-adjusted HbA1c cutoff (children: 73% [67, 78]; adults: 49% [31, 61]; p=0.03). This residual difference may represent underlying biological variation in disease tempo, with children progressing more rapidly once dysglycaemia develops. This may also reflect heterogeneity within the adult cohort.

Among participants positive for multiple autoantibodies, children continued to progress more rapidly than adults even aver accounting for age-related effects on HbA1c. This residual difference is consistent with heterogeneity in disease tempo. Recent histological studies have shown that early life is characterized by numerous small β-cell clusters, which appear to be preferentially lost in type 1 diabetes and are most depleted in children, whereas adults retain a greater proportion of larger islet structures (24). Such age-related differences in β-cell architecture may contribute to faster progression in children and a broader range of, oven slower, progression trajectories in adults aver dysglycaemia develops.

Interpretation of adult risk estimates, particularly when using the higher WHO-defined HbA1c thresholds, must consider reduced statistical power. Application of age-adjusted HbA1c (age-adjusted HbA1c ≥ 5.7% or 39 mmol/mol) and WHO-defined (HbA1c ≥ 6.0% or 42 mmol/mol) cutoffs identified fewer adults with dysglycaemia, widening confidence intervals and limiting sensitivity to detect modest differences in progression risk. Larger adult cohorts with longer follow-up will be required to more precisely define progression patterns within multiple autoantibody positive adults.

HbA1c is not the only measure of glycemia; Stage 2 type 1 diabetes is also identified by OGTT by ADA guidelines(25), while consensus criteria further incorporate continuous glucose monitoring (CGM) measures (4). However, OGTT findings also vary by age, and it is likely that OGTT, like HbA1c, may also require some age adjustment to accurately stage presymptomatic type 1 diabetes. Unfortunately, large population datasets are not available to establish how OGTT cutoffs should be interpreted in children versus adults.

Our study has several limitations. We studied primarily first-degree relatives (93% of adults were first degree relatives). However, this relative status applied to both children and adults in our analysis and several general population screening studies have highlighted progression risk and natural history is similar between relatives and the general population once stage 1 type 1 diabetes is reached (26).

Most participants in our study self-reported ethnicity and race as non-Hispanic white however it is known that HbA1c varies by ancestry, specifically individuals of non-Hispanic and black ethnicity had higher HbA1c for the same glycemia (27), and therefore potential differences by ancestry may need to be addressed. Numbers of adults with multiple ABs, especially with long-term follow-up were relatively low, increasing the size of confidence intervals. Larger cohorts are needed to increase the estimation of the sensitivity and specificity for each HbA1c adjustment approach. Although a 10% increase in HbA1c is considered as part of stage 2 type 1 diabetes criteria, (4), analysis of change in HbA1c in adults was limited by timing and availability of follow-up HbA1c data. Future work will need to test if the 10% HbA1c increase is similarly predictive in adults as children.

We also explored whether implementing an age cut-off could help determine when the increased HbA1c threshold should no longer apply to younger adults (<30 years). Such an approach could facilitate smoother transitions from pediatric to adult care by maintaining consistent diagnostic criteria across services. The choice of 30 years as a provisional cut-off was informed by prior analyses (20,21); however, given the gradual age-related increase in HbA1c across adulthood, this boundary should be viewed as pragmatic rather than definitive, and additional data will be needed to refine the optimal age at which higher thresholds are most appropriate.

A key strength of our findings is their direct clinical relevance. HbA1c is already widely available, inexpensive, and familiar to clinicians globally, unlike many emerging biomarkers. Our work demonstrates that with appropriate thresholds, HbA1c can serve as a practical and effective predictor of progression in AB positive adults. Higher thresholds in adults effectively limit false positives while maintaining a very low progression risk among those below the threshold. This highlights HbA1c as an attractive tool for risk stratification in clinical seungs, facilitating earlier identification of high-risk individuals who may benefit from monitoring or intervention.

Although age-adjusted HbA1c may offer the most physiologically accurate approach, it requires additional computational steps and may be therefore better suited for research contexts rather than clinical seungs. From a translational perspective, clinical practice should balance simplicity with predictive performance. The increased, WHO-defined threshold, HbA1c ≥ 6.0% (42 mmol/mol), provides a pragmatic, readily implementable option for adults, aligning with international standards while improving risk discrimination.

From a clinical perspective, our findings support use of a higher HbA1c threshold (≥6.0% or 42 mmol/mol) to define dysglycaemia in autoantibody-positive adults aged ≥30 years. Although age-adjusted HbA1c accounts for non-glycaemic age effects and performs similarly in risk stratification, its reliance on external reference data limits routine clinical use. In contrast, a fixed HbA1c threshold of 6.0% is simple to implement and aligns adult progression risk more closely with that observed in children and younger adults at the standard 5.7% (39 mmol/mol) cutoff. We therefore recommend reserving age-adjusted HbA1c for research applications and adopting HbA1c ≥6.0% for clinical screening and follow-up of adults aged ≥30 years as screening programs expand.

To conclude, HbA1c is a useful predictor of progression to type 1 diabetes in both children and adults with positive islet autoantibodies. In adults aged ≥ 30 years, the risk of disease progression associated with a higher HbA1c threshold (HbA1c ≥ 6.0% or 42 mmol/mol) aligns more closely with the risk observed in children and adults <30 years at the 5.7% (39 mmol/mol) threshold. In clinical seungs, this supports adoption of an HbA1c threshold of ≥6.0% (42 mmol/mol) for autoantibody-positive adults aged ≥30 years, while age-adjusted HbA1c may be most appropriately applied in research contexts. This age-stratified approach provides a simple, clinically implementable framework for improving dysglycaemia assessment, using thresholds consistent with widely used international diabetes cutoffs for dysglycaemia in type 2 diabetes.

## Data Availability

All data are available through the NIDDK Central Repository via controlled-access request.

## Author Contributions

ELT, NT, LAF, EKS, and RAO conceived the idea and designed the study. ELT analysed the data with assistance from NT, LAF, EKS, and RAO. ELT, NT, LAF, EKS, and RAO interpreted the results. ELT draved the manuscript with assistance from NT, LAF, EKS and RAO. All authors critically revised the manuscript and approved the final version. The corresponding author attests that all listed authors meet authorship criteria and that no others meeting the criteria have been omitted.

