## Supplementary for "Accounting for age-related increases in HbA1c more accurately quantifies risk of Type 1 Diabetes progression in islet autoantibody-positive adults"

### Supplementary Methods:

Variable Definition: HbA1c values were converted from percentage units (%; National Glycohemoglobin Standardization Program, NGSP) to mmol/mol (International Federation of Clinical Chemistry, IFCC) using the standard transformation formula:

**IFCC = (NGSP − 2.15) × 10.929.**

#### Statistical Analysis:

Additional survival analyses applied alternative HbA1c thresholds (e.g. ≥6.0%), age stratification in adults (<30 years vs ≥30 years), and autoantibody status (single vs multiple autoantibodies). If autoantibodies were not drawn at the same visit as HbA1c, the last known autoantibody status was taken.

##### Modeling HbA1c Measurements

The measurement for the participant *i* of the measurement of HbA1c at age *t* is modeled as

$$y_{i}(t)=\beta+f\left( t \right)+\varepsilon_{i},$$

where,

- $\beta$ is the average HbA1c,
- $f\left( t \right)$ representing the natural increase in HbA1c over time. nd we assume that $f\left( t \right)=0$ for all $t\leq18$ assuming that there is no increase in HbA1c for ages 18 or younger.
- $\varepsilon_{i}$ is the random error, assumed to follow $\varepsilon_{i}\sim N\left( 0,\sigma^{2} \right)$ , a

We fitted a linear model, predicting HbA1c by age, in EXTEND so that

$$f\left( t \right)= \alpha\cdot\left( t - 18 \right), if t > 18$$

where, $\alpha$ is estimated from the model developed in EXTEND.

##### **Adjusting HbA1c for TrialNet Participants**

To remove the effect of time from the HbA1c measurements for the TrialNet participants *j* we want to estimate $\tilde{y}_{j}$, given $y_{j}(t)$ is the measured HbA1c in TrialNet, the age *t* of the person :

$$\tilde{y}_{j}=\beta+\varepsilon_{j}$$

One can remark that:

$$\tilde{y}_{j}=y_{j}(t)- f\left( t \right)=y_{j}(t) -\alpha\cdot(t-18)$$

### Supplementary Table and Figures:

Supplementary Table 1

|  | Adults (N=1304) | Children (N=3720) | P-value |
| --- | --- | --- | --- |
| **Age at screening** |  |  | <0.001 |
| Mean (SD) | 34.9 (8.39) | 9.48 (4.07) |  |
| Median [Min, Max] | 36.9 [18.0, 51.2] | 9.34 [1.15, 20.5] |  |
| **Sex** |  |  | <0.001 |
| Female | 842 (64.6%) | 1695 (45.6%) |  |
| Male | 462 (35.4%) | 2025 (54.4%) |  |
| **Ethnicity** (% White) | 1,115 (85.5%) | 3,212 (86.3%) | 0.532 |
| **z_bmi** |  |  | <0.001 |
| Mean (SD) | 1.33 (1.32) | 0.49 (1.32) |  |
| Median [Min, Max] | 1.22 [-2.34, 6.30] | 0.40 [-11.3, 11.7] |  |
| **BMI** |  |  | <0.001 |
| Mean (SD) | 27.5 (6.33) | 18.6 (4.64) |  |
| Median [Min, Max] | 26.0 [16.0, 56.1] | 17.3 [4.82, 95.2] |  |
| **Develop T1D** | 207 (15.9%) | 1386 (37.3%) | <0.001 |
| **HbA1c meeting dysglycaemia**  (HbA1c > 39 mmol/mol or 5.7%) | 110 (8.4%) | 198 (5.3%) | <0.001 |
| **Number of Autoantibodies** |  |  | <0.001 |
| Single | 990 (75.9%) | 1435 (38.6%) |  |
| Multiple | 314 (24.1%) | 2285 (61.4%) |  |
| **GADA positive** | 1118 (85.7%) | 3076 (82.7%) | 0.012 |
| **IA2A positive** | 253 (19.4%) | 1619 (43.5%) | <0.001 |
| **IAA positive** | 248 (19.0%) | 1982 (53.3%) | <0.001 |

Baseline characteristics of adult (≥ 18 years at screening) and children (<18 years at screening) relatives enrolled in the TrialNet Pathway to Prevention study. Data are presented as mean (SD) and median [minimum and maximum] for continuous variables, and percentage for categorical variables. Differences between adults and children are indicated through p-values.

Supplementary Table 2

|  | Single Autoantibody Positive (N=990) | Multiple Autoantibody Positive (N=314) | P-value |
| --- | --- | --- | --- |
| **Age at screening** |  |  | 0.005 |
| Mean (SD) | 35.9 (7.86) | 31.8 (9.23) |  |
| Median [Min, Max] | 37.5 [18.0, 51.2] | 33.6 [18.0, 49.5] |  |
| **Sex** |  |  | <0.001 |
| Female | 673 (68.0%) | 169 (53.8%) |  |
| Male | 317 (32.0%) | 145 (46.2%) |  |
| **Ethnicity** (% White) | 847 (85.6%) | 268 (85.4%) | 0.815 |
| **z_bmi** |  |  | 0.749 |
| Mean (SD) | 1.35 (1.31) | 1.28 (1.38) |  |
| Median [Min, Max] | 1.21 [-2.34, 6.30] | 1.28 [-1.84, 5.68] |  |
| **BMI** |  |  | 0.954 |
| Mean (SD) | 27.5 (6.33) | 27.4 (6.36) |  |
| Median [Min, Max] | 25.9 [16.0, 56.1] | 26.3 [16.8, 53.6] |  |
| **Develop T1D** | 99 (10.0%) | 108 (34.4%) | <0.001 |
| **HbA1c meeting dysglycaemia**  (HbA1c > 39 mmol/mol or 5.7%) | 70 (7.1%) | 40 (12.7%) | <0.001 |
| **Number of Autoantibodies** |  |  | <0.001 |
| 1 | 990 (100%) | 0 (0%) |  |
| 2 | 0 (0%) | 314 (100%) |  |
| **GADA positive** | 812 (82.0%) | 306 (97.5%) | <0.001 |
| **IA2A positive** | 35 (3.5%) | 218 (69.4%) | <0.001 |
| **IAA positive** | 158 (16.0%) | 90 (28.7%) | <0.001 |

Baseline characteristics of adult (≥ 18 years at screening) enrolled in the TrialNet Pathway to Prevention study stratified by their autoantibody status at screening (single autoantibody positive vs. multiple autoantibody positive. Data are presented as mean (SD) and median [minimum and maximum] for continuous variables, and percentage for categorical variables. Differences between single and multiple autoantibody positive adults are indicated through p-values.

Supplementary Table 3

| Variable | Exeter 10,000 (EXTEND) |
| --- | --- |
| N | 4,298 |
| Sex (% Female) | 2,815 (65%) |
| Age [years] (SD) | 43.1 (10.3) |
| Ethnicity (% White) | 4,180 (97%) |
| BMI (SD) | 26.2 (4.9) |
| HbA1c [mmol/mol] (SD) | 36.5 (3.7) |

Characteristics of the population in Exeter 10,000 (EXTEND). Data are presented as mean (SD) for continuous variables, and n (%) for categorical variables.

Supplementary Table 4

| Age | Age - Adjusted HbA1c | | Measured HbA1c (mmol/mol) | |
| --- | --- | --- | --- | --- |
|  | mmol/mol | % | mmol/mol | % |
| 18 | 39 | 5.7 | 39 | 5.7 |
| 20 | 39 | 5.7 | 39 | 5.7 |
| 25 | 39 | 5.7 | 40 | 5.8 |
| 30 | 39 | 5.7 | 41 | 5.9 |
| 35 | 39 | 5.7 | 41 | 5.9 |
| 40 | 39 | 5.7 | 42 | 6.0 |
| 45 | 39 | 5.7 | 43 | 6.1 |
| 50 | 39 | 5.7 | 43 | 6.1 |
| 55 | 39 | 5.7 | 44 | 6.2 |

Age-adjusted rounded to the nearest mmol/mol HbA1c equivalents to measured HbA1c by age. The table shows the measured HbA1c values (by 5-year age intervals) that correspond to an age-adjusted HbA1c of 39 mmol/mol or 5.7%. For example, adults aged ≥ 40 years required a measured HbA1c of 42 mmol/mol (6.0%) or higher to be equivalent to an age-adjusted HbA1c of 39 mmol/mol (5.7%).

Supplementary Figure 1

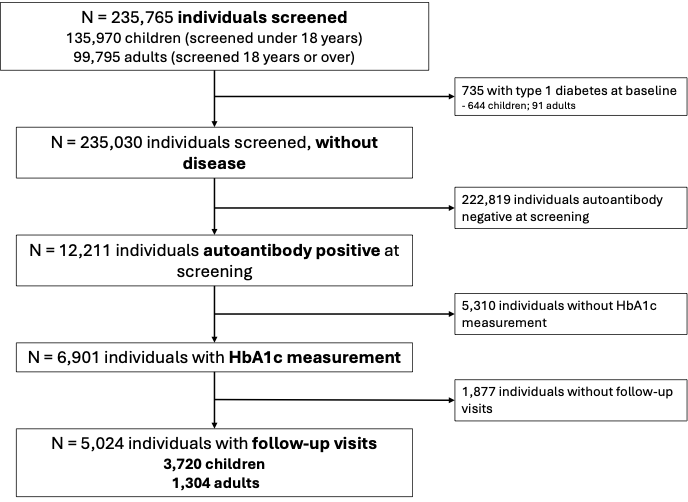

Consort diagram demonstrating the TrialNet participants included in current analysis. Of the 235,765 individuals screened, 230,741 individuals were excluded due to type 1 diabetes identified at screening/baseline (n=735), individuals are autoantibody negative at screening/baseline (n=222,819), individuals without HbA1c measurements (n=5,310), and indviduals without follow-up (n=1,877). The final analysis included 5,024 individuals with follow-up visits, comprising 3,720 children and 1,304 adults.

Supplementary Figure 2

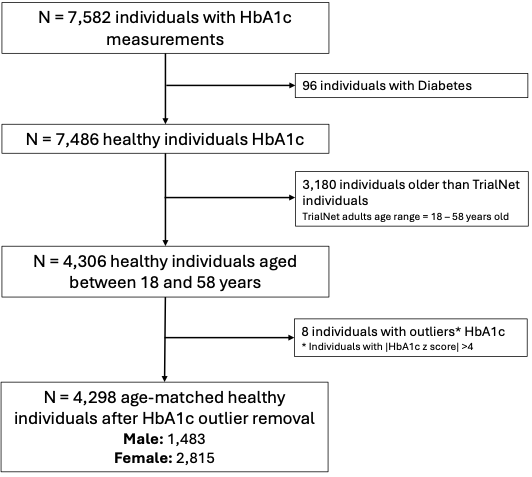

Consort diagram demonstrating the Exeter 10,00 Biobank (EXTEND) participants included in current analysis. Of the 7,582 individuals with HbA1c measurements in EXTEND, 3,284 individuals were excluded due to being identified with diabetes (n=96), individuals were older than those in TrialNet (n=3,180), and individuals with a |HbA1c z-score| of greater than 4 (n=8). Assuming a normal distribution, we would expect 1 in 10,000 people to have a |z-score| > 4 and therefore as we have less than this number of individuals, we have assumed these individuals (n=8) to be outliers in our distribution and therefore can act as influential points in the modelling process.

Supplementary Figure 3

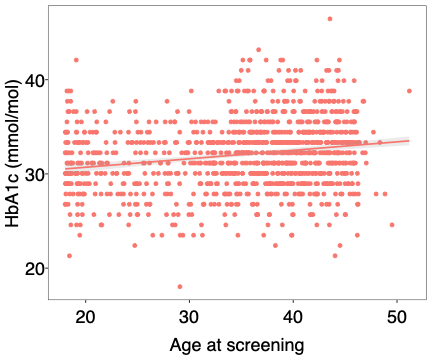

Linear Regression model and scatter plot of HbA1c values (mmol/mol) by age (years) in the adults followed in the TrialNet Pathway to prevention study. Each dot represents an individual participant. The solid line indicates the fitted linear regression model and the shaded band around the regression line represents the 95% confidence interval. In TrialNet, the effect of age when predicting HbA1c in nonprogressors (those that remain type 1 diabetes free over the follow-up) is 0.0134 [95% CI 0.063, 0.116] (p<0.001).

Supplementary Figure 4

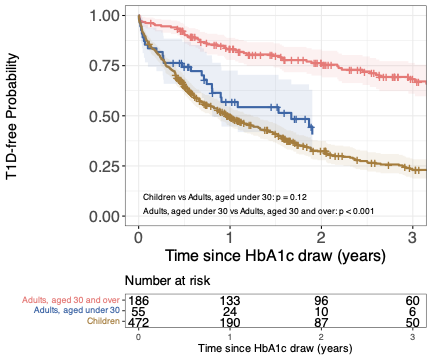

Kaplan-Meier (survival) graphs of TrialNet participants stratified by age (Children, aged under 18 years, Adults aged under 30 years, and Adults aged 30 years and over) from when individuals are identified with dysglycaemia, defined by HbA1c ≥ 39 mmol/mol. Adults, aged 30 years and over compared to adults, aged under 30 years have significantly lower risk of type 1 diabetes (p<0.001), however adults, aged under 30 years to children have similar risk of type 1 diabetes (p=0.12). Survival curves are truncated when they contain 10 individuals or less.

Supplementary Figure 5
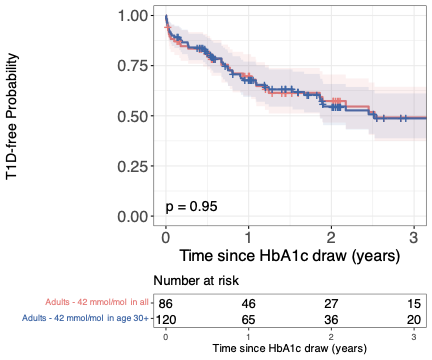

Kaplan-Meier (survival) graphs comparing two approaches: *“Adults – 42 mmol/mol in all”* uses the higher cut-off (HbA1c ≥ 42 mmol/mol) in all adults to identify dysglycaemia, : *“Adults – 42 mmol/mol in age 30+”* uses the higher cut-off (HbA1c ≥ 42 mmol/mol) in adults aged 30 years and above and the original cut-off (HbA1c ≥ 39 mmol/mol) in adults aged under 30 years to identify dysglycaemia and for adults aged less than 30 years, the original threshold, proposed in the consensus guidelines (HbA1c ≥ 39 mmol/mol) is used to identify dysglycaemia. There is no significant difference the risk of T1D between the cohorts (p=0.95), similarly identifying adults with a 50% risk of T1D in 3 years.

Supplementary Figure 6

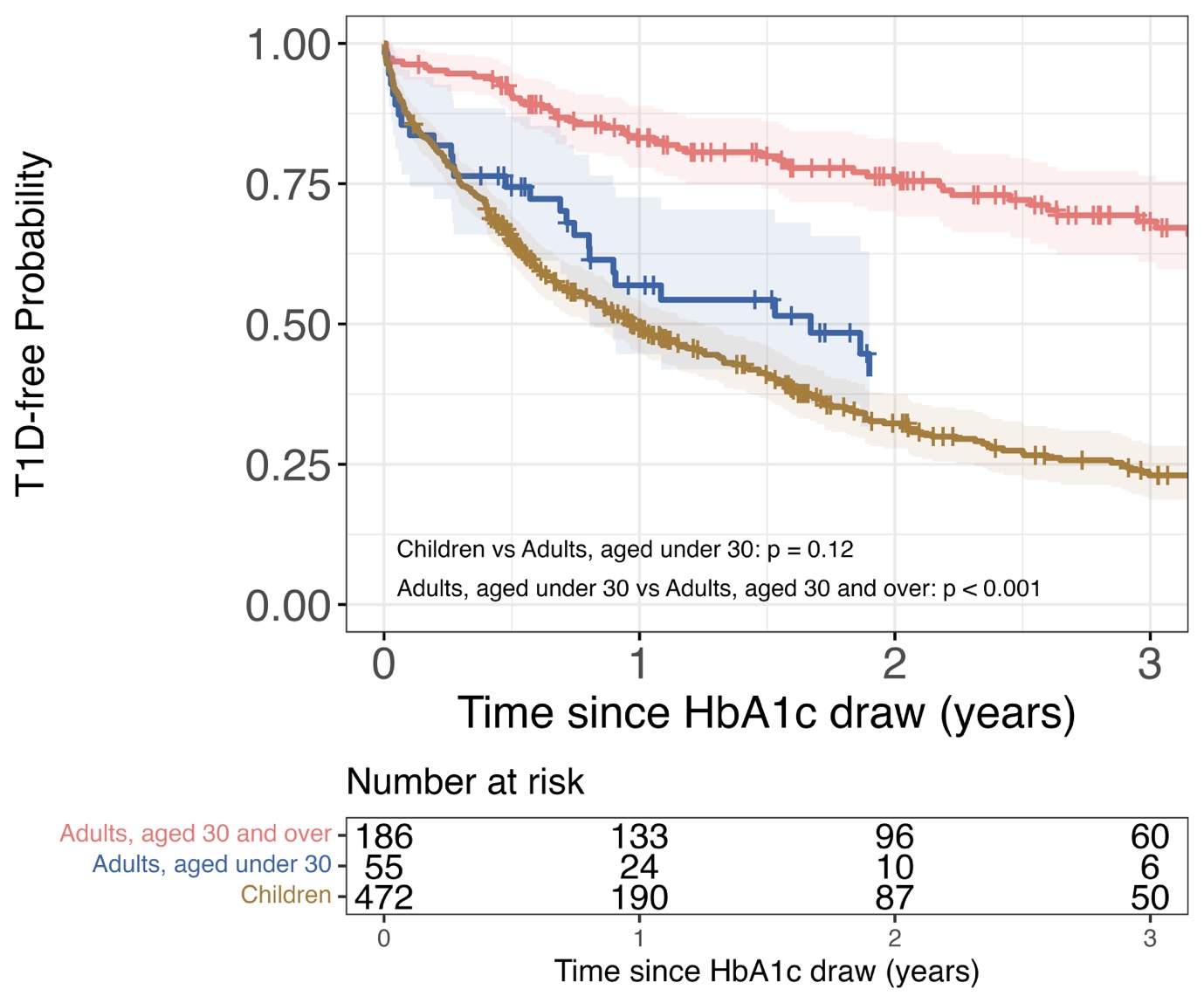

Kaplan-Meier (survival) graphs comparing two risk of adults using two different approaches *“Adults – 42 mmol/mol in all”* or *“Adults – 42 mmol/mol in age 30+”* to the risk of children. *“Adults – 42 mmol/mol in all”* uses the higher cut-off (HbA1c ≥ 42 mmol/mol) in all adults to identify dysglycaemia, *“Adults – 42 mmol/mol in age 30+”* uses the higher cut-off (HbA1c ≥ 42 mmol/mol) in only adults aged 30 years and above to identify dysglycaemia and for adults aged less than 30 years, the original threshold, proposed in the consensus guidelines (HbA1c ≥ 39 mmol/mol) is used to identify dysglycaemia. Children are identified using the original threshold (HbA1c ≥ 39 mmol/mol) to identify dysglycaemia. Children consistently have a higher risk of T1D than adults, regardless of the approach to identify dysglycaemia (p<0.001 for both, *Children* vs *Adults – 42 mmol/mol in all*, and *Children* vs *Adults – 42 mmol/mol in age 30+*.

Supplementary Figure 7
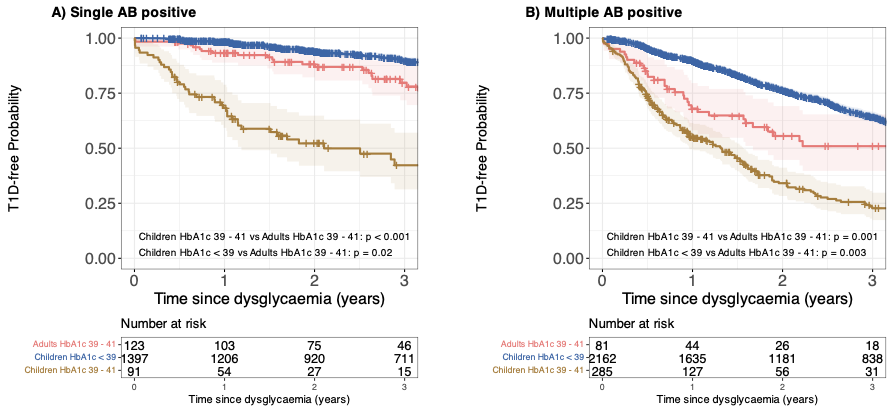

Kaplan-Meier (survival) graphs of TrialNet “low risk” Adults and Children stratified by autoantibody status at HbA1c, categorised into Children less than 39 mmol/mol, Children between 39 mmol/mol and 41 mmol/mol, and Adults between 39 mmol/mol and 41 mmol/mol. Survival curves are truncated when they contain 10 individuals or less.

Supplementary Figure 8

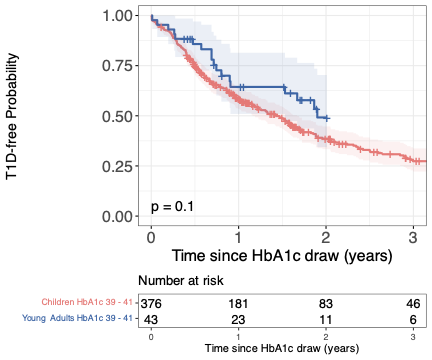

Kaplan-Meier (survival) graphs of TrialNet young adults (aged under 30 years) to Children with HbA1c between 39 mmol/mol to 41 mmol/mol. Survival curves are truncated when they contain 10 individuals or less.
